# Inpatient diagnostic odysseys in rare diseases: a nationwide audited Orphanet–ICD-10–DRG/GRD-IR analysis in Chile, 2019–2024

**DOI:** 10.64898/2026.05.01.26352213

**Authors:** Germán Gómez-Vargas, Gabriela Repetto, Loreto Bravo, Carla C. Castillo-Laborde, Iris Delgado, María Isabel Matute Willemsen

## Abstract

**Background:** Rare diseases (RD; *enfermedades poco frecuentes*, EPoF, in Chilean policy terminology) collectively affect 3.5–5.9% of the population and are associated with long diagnostic trajectories. Chile lacks a reproducible national operational definition for identifying RD in administrative hospital data.

**Methods:** We conducted a retrospective observational analysis of Chilean GRD-IR events (IR-29301 version) for 2019–2024 released through FONASA Datos Abiertos, covering hospital discharges and major ambulatory surgery reported by 72 public establishments for FONASA-covered persons. The canonical analytical cohort contained 5,779,482 DRG events in 4,027,921 linked patients. We constructed a Chilean Orphanet–ICD-10 homologation and audited it through an agentic human-in-the-loop pipeline, yielding a conservative RD operational catalogue (434 final ICD-10 codes in the `KEEP + MAP_TO_SPECIFIC_ORPHA` scenario). RD-coded DRG events were labeled as observed inpatient odysseys when at least one prior DRG event existed for the same patient. We quantified prior events, DRG-observed inpatient trajectory time, nonspecific prior diagnoses, DRG weight, and bridge-code associations. Bridge-code enrichment was estimated using patient-level Fisher exact tests with Benjamini–Hochberg false-discovery correction; event-level estimates were retained as sensitivity analyses.

**Results:** The audited conservative catalogue identified 55,284 primary-diagnosis RD-coded DRG events in 45,784 patients and 374,866 RD-coded events in any diagnostic field. We characterized 63,685 observed inpatient odyssey cases in 25,648 unique patients across 371 audited RD ICD-10 codes. Median DRG-observed inpatient trajectory time to RD-coded diagnosis was 241 days, and mean prior events per odyssey was 8.1. Bridge-code analysis identified 616 associations with support ≥ 10 patients and 390 with *q <* 0.05; 350 significant associations were no-same-code administrative trajectory signals. These signals varied in interpretation, including clinically plausible precursors, diagnostic refinement, and care-process bridges. The Odyssey Index reordered conditions relative to raw prior-event counts, separating high-volume entities from stronger trajectory signatures.

**Conclusions:** To our knowledge, we provide the first nationwide audited and reproducible characterization of inpatient RD diagnostic odysseys in Latin America using administrative hospital data. The framework supports trajectory surveillance, registry design, quality-control analyses, and prioritization of candidate signals for prospective clinical validation under Chile’s Law 21,743. Bridge-code associations should be interpreted as statistically enriched administrative signals, not as validated causal or clinical pathways.

Graphical abstract. Updated canonical FONASA DRG/GRD-IR 2019–2024 cohort, audited RD catalogue, odyssey cohort, and bridge-code signal summary.

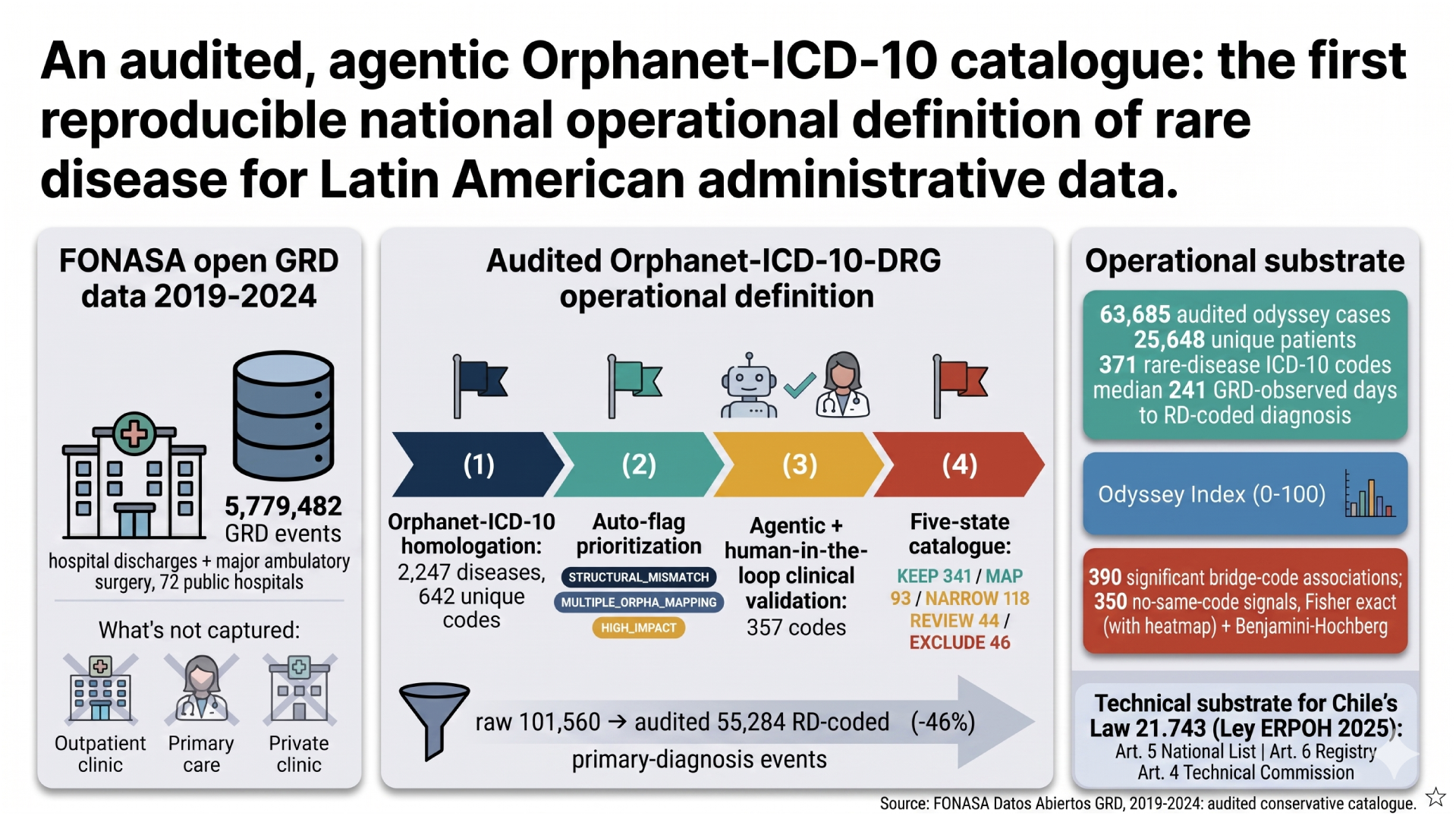

## 1 Introduction

Rare diseases (RD) are individually rare but collectively affect 3.5–5.9% of the population [8]. More than 6,000 distinct entities have been recognized; ∼80% are genetic and ∼70% have pediatric onset [8, 5]. Their chronic and disabling nature creates a substantial burden on health systems, families, and labor participation [13, 4].

The pan-European Rare Barometer survey (Faye et al. 2024; n = 6,507 in 41 countries) reported a median total diagnostic time of **4.7 years**, with 56% of patients diagnosed more than 6 months after first medical contact [5]. The diagnostic odyssey has been qualitatively well described, including in Chile by Cabieses, Obach, Roberts and Repetto (2025) [3], but it remains poorly quantified at the population level, particularly in Latin America.

Population-wide registries enable temporal trajectory mining [11, 12]. Algorithmic search (Ediae et al. *ThinkRare* 2025) and agentic LLM systems with traceable reasoning (Zhao et al. *DeepRare* 2026) are emerging as diagnostic copilots [6, 7]. In Latin America, however, this approach has not been systematically applied to RD using administrative data.

In May 2025, Chile enacted **Law 21,743 – Law ERPOH**, the country’s first explicit national regulatory framework for rare, orphan or infrequent diseases [14]. The law (a) formally defines a rare disease as one with prevalence below 1 per 2,000 inhabitants (Art. 2); (b) creates a *Technical Advisory Commission on Rare Diseases* (Art. 4); (c) instructs the Ministry of Health to maintain an up-to-date *national list of rare diseases* with biennial validity (Art. 5); and (d) creates a *National Registry of persons with rare diseases* (Art. 6), regulated by a technical norm (Decree 1 Exento, Chilean Ministry of Health [MINSAL], Feb 2026). This framework creates an institutional need for reproducible operational definitions, registry-compatible analytics and transparent evidence-generation procedures. The present work addresses the administrative-data component of that need, without replacing the prevalence assessment, ministerial validation, physician-confirmed diagnosis or consent-based enrolment required for formal implementation. Law 21,743 now provides the central disease-specific regulatory framework for Chilean rare-disease policy, while Law 20,850 (*Ricarte Soto*) [15] continues to govern high-cost coverage for a subset of conditions. These legal developments are preceded and accompanied by earlier and parallel national efforts, including the country position statement [2], Repetto’s editorial perspective [4], the Goic et al. proposal for a National Plan for Rare Diseases [9], and the institutional development of the Ministry of Health’s National Office for Complex Chronic Conditions and Rare Diseases (*Oficina Nacional de Condiciones Crónicas Complejas y Enfermedades Poco Frecuentes*), which has also contributed to national planning in this field [10].

Avila and Martínez characterized rare-disease mortality in Chile from 2002 to 2017 using mortality records from the Department of Health Statistics and Information (*Departamento de Estadísticas e Información de Salud*, DEIS) of the Chilean Ministry of Health, and explicitly noted the absence of a Chilean operational definition, reusing a Colombian homologation [1]. The complementary morbidity side—longitudinal inpatient diagnostic trajectories based on a Chilean-built, audited Orphanet–ICD-10–DRG/GRD-IR catalogue—has not been developed at national scale.

We aim to (i) build an audited Orphanet–ICD-10–DRG/GRD-IR operational catalogue for identifying RD-coded DRG events in Chilean administrative data; (ii) characterize, at national scale in Latin America, the observed inpatient diagnostic-trajectory signature; and (iii) release a reproducible agentic auditing pipeline with human-in-the-loop, in line with Line 1 of the Chilean National Plan and complementary to ThinkRare and DeepRare.

## 2 Scientific contribution

1. **To our knowledge, the first audited operational RD catalogue for administrative data in Latin America.** We publish a Chilean Orphanet–ICD-10 homologation (2,247 diseases, 99.9% mapping success) audited via prioritization by epidemiological impact and mapping risk; five-state agentic classification (KEEP, MAP, NARROW, REVIEW, EXCLUDE); and human clinical review per code (357 priority codes validated). Output: a conservative catalogue (434 codes, 701 rows) and a sensitivity catalogue, both open. This addresses an explicit gap noted by Avila & Martínez (2022).
2. **To our knowledge, the first nationwide systematic characterization of inpatient RD diagnostic odysseys in Latin America.** Unlike Avila 2022, which covered cross-sectional mortality, this work describes the longitudinal pre-diagnostic trajectory: 63,685 odyssey cases in 25,648 unique patients over 371 audited rare-disease ICD-10 codes, with metrics for time, complexity and inpatient resource use.
3. **Quantitative identification of bridge codes with false-discovery control.** We formally define a *bridge code* as the immediately prior primary ICD-10 code before an RD-coded DRG event and quantify the association at the patient level (primary) and event level (sensitivity) via Fisher exact + Benjamini–Hochberg, with Haldane–Anscombe correction in zero cells. We report 350 *no same-code* significant associations (*q <* 0.05, support ≥ 10 patients), including candidate administrative trajectory signals such as N04.9 → N04.8, I67.1 → Q28.2, and D69.4 → D69.3.
4. **Odyssey Index: composite metric robust to volume.** A 0–100 index integrating prior events, observed inpatient trajectory time, fraction of nonspecific prior diagnoses, and DRG weight. The index separates stronger trajectory signatures from sheer high volume, illustrated by the updated dissociation between high-volume codes such as E76.2 and lower-volume, high-index trajectory signatures in Panel C.
5. **National estimation of pre-diagnostic resource-use complexity.** We use IR_29301_PESO as a proxy for complexity/resource use and report cumulative pre-diagnostic inpatient burden per condition.
6. **Reproducible agentic pipeline published.** Audit plan, prioritization workpacks, clinical-validation template, the `validacion-clinica-epof` skill (with human-in-the-loop), and reproduction scripts are released. The core scripts are:
  - `construir_workpacks.py`: builds prioritized code-review workpacks from national, hospital and mapping-risk signals.
  - `proponer_decisiones_iniciales.py`: assigns the first automatic audit decision per ICD-10 code.
  - `aplicar_catalogo_auditado.py:` applies validated decisions and builds the conservative analysis catalogue.
  - `generar_paquetes_revision_clinica.py:` creates per-code clinical review packages for validation.
  - `consolidar_validacion_clinica.py:` merges clinical-review outputs into the master de-cision table.
  - `regenerar_figura_principal_auditada.py:` regenerates the audited main figure and panel-level outputs.
  This sits alongside ThinkRare (rule-based individual EMR search) and DeepRare (agentic LLM differential): our contribution is the *population-level audited infrastructure* on which such individual copilots can be anchored.
7. **Policy relevance for the implementation of Law 21,743 (Law ERPOH).** The audited operational catalogue, bridge-code analysis and Odyssey Index provide a reproducible technical substrate that can inform the implementation of Law 21,743 (Law ERPOH). For **Art. 5 – the national rare-disease list**, the Orphanet–ICD-10 audit offers a transparent starting point for translating rare-disease entities into administrative coding rules, subject to prevalence assessment, expert deliberation and ministerial validation. For **Art. 6 – National Registry of persons with rare diseases**, the audited catalogue may support registry design, epidemiological monitoring, case-finding support and quality-control analyses, without replacing physician-confirmed diagnosis, consent-based enrolment or the legal requirements of the registry. For **Art. 4 – Technical Advisory Commission**, the agentic human-in-the-loop pipeline provides a methodological framework that could be reviewed, adapted and governed by the Commission. Complementarily, the work may support Law 20,850 (Ricarte Soto) coverage monitoring for high-cost RD, sentinel-hospital validation pilots, referral-network design and the quantitative operationalisation of Line 1 of the proposed National Plan for Rare Diseases.

## 3 Methods

### 3.1 Study design

Retrospective observational study over GRD-IR events coded in Chile’s Diagnosis-Related Groups system (DRG; Chilean GRD-IR, version IR-29301) released through FONASA Datos Abiertos for 2019–2024. The analytical cohort covers hospital discharges and major ambulatory surgery reported by 72 public establishments for persons covered by the National Health Fund (*Fondo Nacional de Salud*, FONASA). Cross-checking the observed facility codes against the establishment dictionary showed that private clinics listed in the dictionary contributed zero events to this 2019–2024 analytical cohort. The data should therefore be interpreted as a FONASA-covered public inpatient administrative cohort, not as a census of all Chilean residents, all FONASA activity, or private-sector care.

### 3.2 The DRG/GRD-IR system: scope, structure, and structural limits

#### What DRG/GRD-IR captures

DRGs classify *closed* hospital episodes into iso-resource categories using diagnoses, procedures, age, sex and outcome. Chile uses the International Refined GRD-IR IR-29301 version, locally calibrated; FONASA publishes the open DRG/GRD-IR files used here. Each record includes an anonymized patient identifier (allowing longitudinal linkage), demographics, reporting establishment and health service, admission/discharge dates, primary (DIAGNOSTICO1) and secondary ICD-10 diagnoses, primary/secondary procedures, the assigned GRD-IR code and weight (`IR_29301_PESO`, a complexity proxy), discharge type, and length of stay. Although the establishment dictionary includes private clinics, the 72 observed facilities in this analytical cohort were public hospitals, public hospital complexes, or national institutes; private clinics such as Clínica Alemana, Clínica Las Condes, and Clínica Santa María had no observed events in the cohort. Facility type therefore could not be used as a public/private stratification variable in the present estimates.

#### What DRG/GRD-IR does not capture

- Outpatient specialty care of any kind (consultations, controls, telemedicine).
- Primary care and ambulatory urgent-care services, including Family Health Centers (*Centros de Salud Familiar*, CESFAM), Primary Emergency Care Services (*Servicios de Atención Primaria de Urgencia*, SAPU), and High-Resolution Emergency Primary Care Services (*Servicios de Alta Resolutividad*, SAR), as well as preventive programs and ambulatory diagnostic tests.
- Emergencies that do not result in hospitalization.
- Private outpatient care and hospitalizations financed outside FONASA.
- Private -sector FONASA hospital activity, including activity that could occur through *Modalidad Libre Elección* or *Ley de Urgencia* mechanisms, is absent from the observed 2019–2024 cohort despite private clinics appearing in the facility dictionary.
- Laboratory results, imaging reports, genetic panels, exome/genome sequencing.
- Outpatient pharmacotherapy (including high-cost drugs covered by Law 20,850 when not associated with hospitalization).
- Non-structured clinical data (epicrisis narrative, chief complaint, evolution).
- Family history, pedigree, and genetic counseling.

#### Structural implications.

DRG/GRD-IR captures only the *inpatient* portion of the trajectory for FONASA-covered persons whose episodes appear in the public reporting establishments. The full odyssey typically begins in primary or specialty outpatient care years before the first hospitalization; therefore, all times and counts here are a *conservative lower bound on the total odyssey*. A patient diagnosed elsewhere in the system, treated entirely in an ISAPRE/private circuit, or hospitalized through private-sector FONASA pathways not represented in these files is invisible to this analysis. This is a major structural, not temporal, limitation: it cannot be resolved by extending the observation window. Resolution requires linkage with primary care, ambulatory emergency, FONASA insurance and claims records, and private-sector records, which is currently not feasible from public data alone.

### 3.3 Auxiliary sources

Orphanet rare-disease catalogue and ICD-10 mappings (https://www.orpha.net); Colombian homologation used by Avila & Martínez (2022) as a comparator; FONASA Datos Abiertos DRG/GRD-IR source files and hospital identifiers; and the Chilean Orphanet–ICD-10 catalogue built by our team in July 2025 and audited in 2026.

### 3.4 Construction of the Chilean Orphanet–ICD-10 homologation

Text-and-code matching procedure between Orphanet entity names/codes and the Chilean official ICD-10 coding, with match types (exact, partial, observational). Final coverage: 2,247 diseases mapped (99.9%). The frozen source catalogue and audited outputs are listed in the Data and code availability section.

### 3.5 Catalogue auditing (agentic + human-in-the-loop)

1. Freezing of the original catalogue and reference national/hospital counts.
2. Automatic flag generation:

- `STRUCTURAL_MISMATCH`
- `NO_ORPHA_ICD_EVIDENCE`
- `MULTIPLE_ORPHA_MAPPING`
- `HIGH_IMPACT_NATIONAL`
- HIGH_IMPACT_FACILITY
3. Prioritization of **642 codes** by impact and mapping risk.
4. Initial automatic decision matrix.
5. Per-code clinical-review package generation.
6. Clinical validation of **357 prioritized codes** by a specialized clinical agent with human-in-the-loop (`skill validacion-clinica-epof`), guided by Orphanet evidence and pre-agreed clinical rules.
7. Final catalogue decisions were treated as human-in-the-loop outputs: the agent generated structured evidence and recommendations, but final inclusion/exclusion decisions were reviewed and accepted by the clinical team before analysis.
8. Consolidation in `decisiones_por_codigo_validado.csv` and release of the audited catalogue.

Final states: KEEP (341), MAP_TO_SPECIFIC_ORPHA (93), NARROW_DEFINITION (118), REVIEW_CLINICAL (44), EXCLUDE (46).

Scenarios: *original*; *audited_conservative* (KEEP + MAP_TO_SPECIFIC_ORPHA, **434 codes / 701 rows**, primary scenario); *sensitivity_no_exclude* (excludes only EXCLUDE; 596 codes / 2,135 rows, plausible upper bound).

### 3.6 Operational definition of diagnostic odyssey

Let *D_p_* = {(*e*_1_*, t*_1_*, c*_1_), …, (*e_n_, t_n_, c_n_*)} be the ordered sequence of DRG events for patient *p*, where *e_i_* is the event index, *t_i_* is the admission date and *c_i_* is the set of ICD-10 codes recorded in the primary or secondary diagnostic fields. Let 𝒞_audit_ denote the audited conservative RD operational catalogue (codes with decision ∈ {KEEP, MAP_TO_SPECIFIC_ORPHA}). A DRG event *e_i_*of patient *p* is labelled an *observed inpatient RD odyssey case* when:

1. *c_i_* ∩ 𝒞_audit_ ≠ ∅ (audited rare-disease coding rule in any diagnostic field); and
2. *i >* 1, i.e., at least one prior DRG event (*e_j_, t_j_, c_j_*) with *t_j_ < t_i_* exists for the same patient, regardless of reporting hospital.

For each odyssey case we compute: (a) prior event count *k_i_* = *i* − 1; (b) DRG-observed inpatient trajectory time to RD-coded diagnosis *τ_i_* = *t_i_* − *t*_1_; (c) bridge code *b_i_* as the primary ICD-10 code of the immediately previous DRG event; (d) fraction of nonspecific prior diagnoses 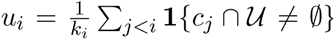, where 𝒰 is a pre-agreed list of generic/unspecified ICD-10 codes (Z-codes for aftercare, R-symptom codes, “unspecified” chapter-end codes); (e) cumulative pre-RD DRG weight 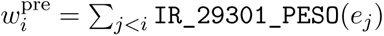; and (f) DRG weight of the RD-coded event itself, 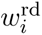.

Applying this definition to the canonical GRD-IR cohort 2019–2024 (*N* = 5,779,482 analyzed events in 4,027,921 linked patients) yielded **63,685 odyssey cases in 25,648 unique patients** spanning **371 audited RD ICD-10 codes**, with median DRG-observed inpatient trajectory time to RD-coded diagnosis 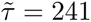 days, mean prior events 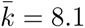, and average DRG weight per RD-coded event 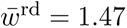. This estimate is a conservative lower bound on the full diagnostic odyssey because outpatient, primary-care, emergency-without-admission, private outpatient trajectories, and private-sector FONASA hospital activity are not captured.

We also computed a stricter *first-late-RD* variant for facility-level feasibility analysis: a patient is in the strict subset only if the first audited RD code in their entire trajectory satisfies *τ* ≥ 30 days; this excludes incidental rare-disease codings adjacent to the index admission.

### 3.7 Bridge codes (administrative trajectory associations)

Cohort: audited odyssey cases (63,685 cases in 25,648 unique patients).

#### Primary analysis – patient unit

For each pair (bridge *c*, RD-coded diagnosis *r*), patient-level 2 × 2 table:

| | Bridge $c$ present | Bridge $c$ absent |
| --- | --- | --- |
| RD code $r$ present | $a$ | $b$ |
| RD code $r$ absent | $c$ | $d$ |

where presence is at the patient level over all audited odyssey events. Support is `pair_patients` (unique patients with the direct *c* → *r* pair, i.e., *c* immediately preceding the RD-coded DRG event). The patient-level analysis was chosen as primary to reduce inflation from repeated events by the same individual. Event-level estimates were retained as sensitivity analyses to evaluate robustness at the case/event level.

#### Sensitivity analysis – case/event unit

The same procedure was replicated at the event unit (one odyssey case per RD-coded DRG event) and tabulated separately; Figure 2D displays the patient-unit primary analysis.

**Figure 1.**
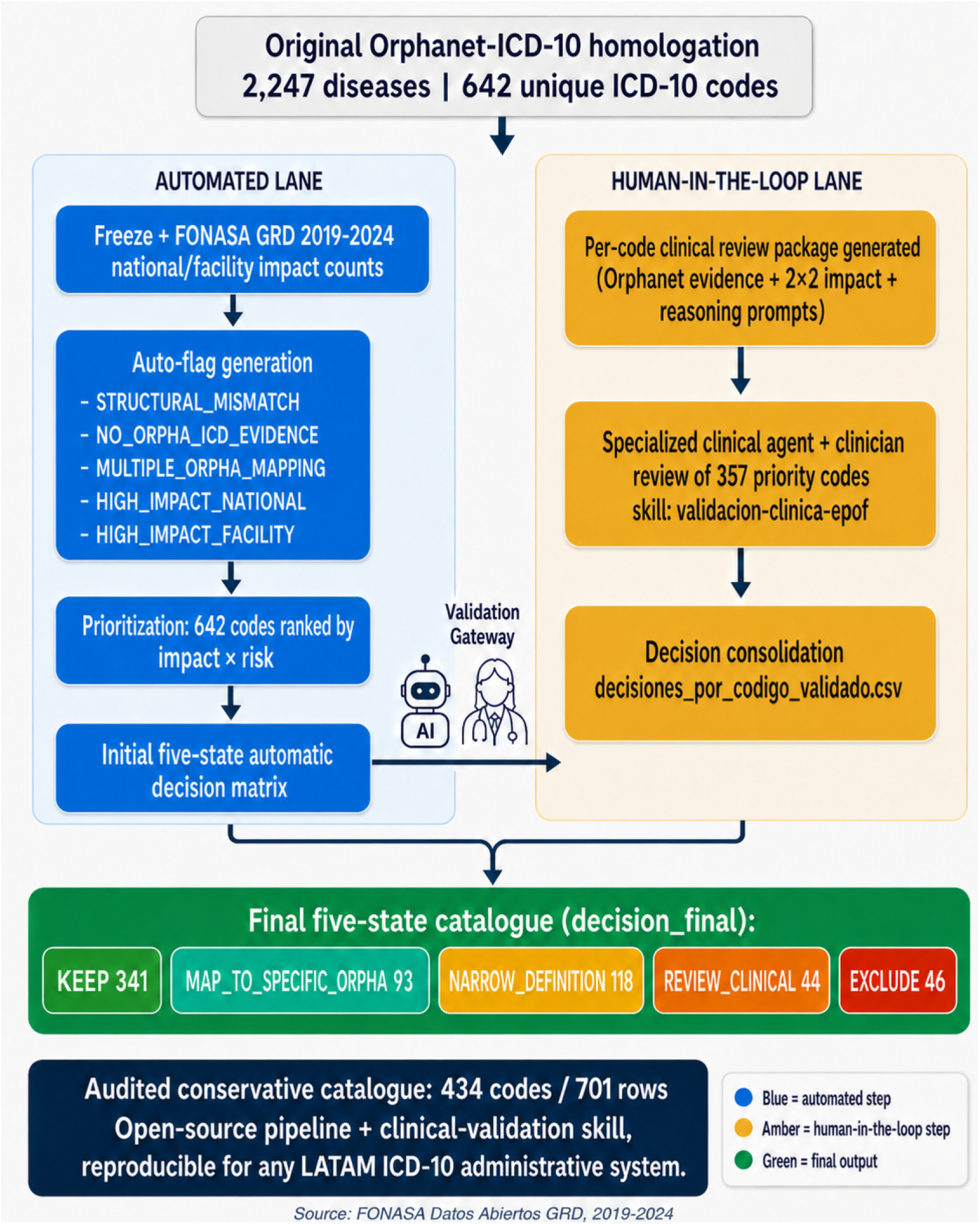
Agentic + human-in-the-loop catalogue auditing pipeline. Two parallel lanes are shown. The *automated lane* (blue) freezes the original Orphanet–ICD-10 homologation (2,247 diseases / 642 unique ICD-10 codes), generates five auto-flags for structural and impact risk (STRUCTURAL_MISMATCH, NO_ORPHA_ICD_EVIDENCE, MULTIPLE_ORPHA_MAPPING, HIGH_IMPACT_NATIONAL, HIGH_IMPACT_FACILITY), prioritises the 642 codes by impact × risk and produces an initial five-state automatic decision matrix. A validation gateway hands off priority cases to the *human-in-the-loop lane* (amber): a per-code clinical review package is generated (Orphanet evidence, 2 × 2 impact, reasoning prompts) and a specialised clinical agent plus a clinician jointly review 357 priority codes through the validacion-clinica-epof skill. Decisions are consolidated in decisiones_por_codigo_validado.csv. The output node carries the final five-state decision_final catalogue: **KEEP 341, MAP_TO_SPECIFIC_ORPHA 93, NAR-ROW_DEFINITION 118, REVIEW_CLINICAL 44, EXCLUDE 46**. The audited conservative catalogue used for the primary analysis is **434 codes / 701 rows**. The full pipeline (skill + scripts) is open-source and adaptable to Latin American ICD-10 administrative systems.

**Figure 2.**
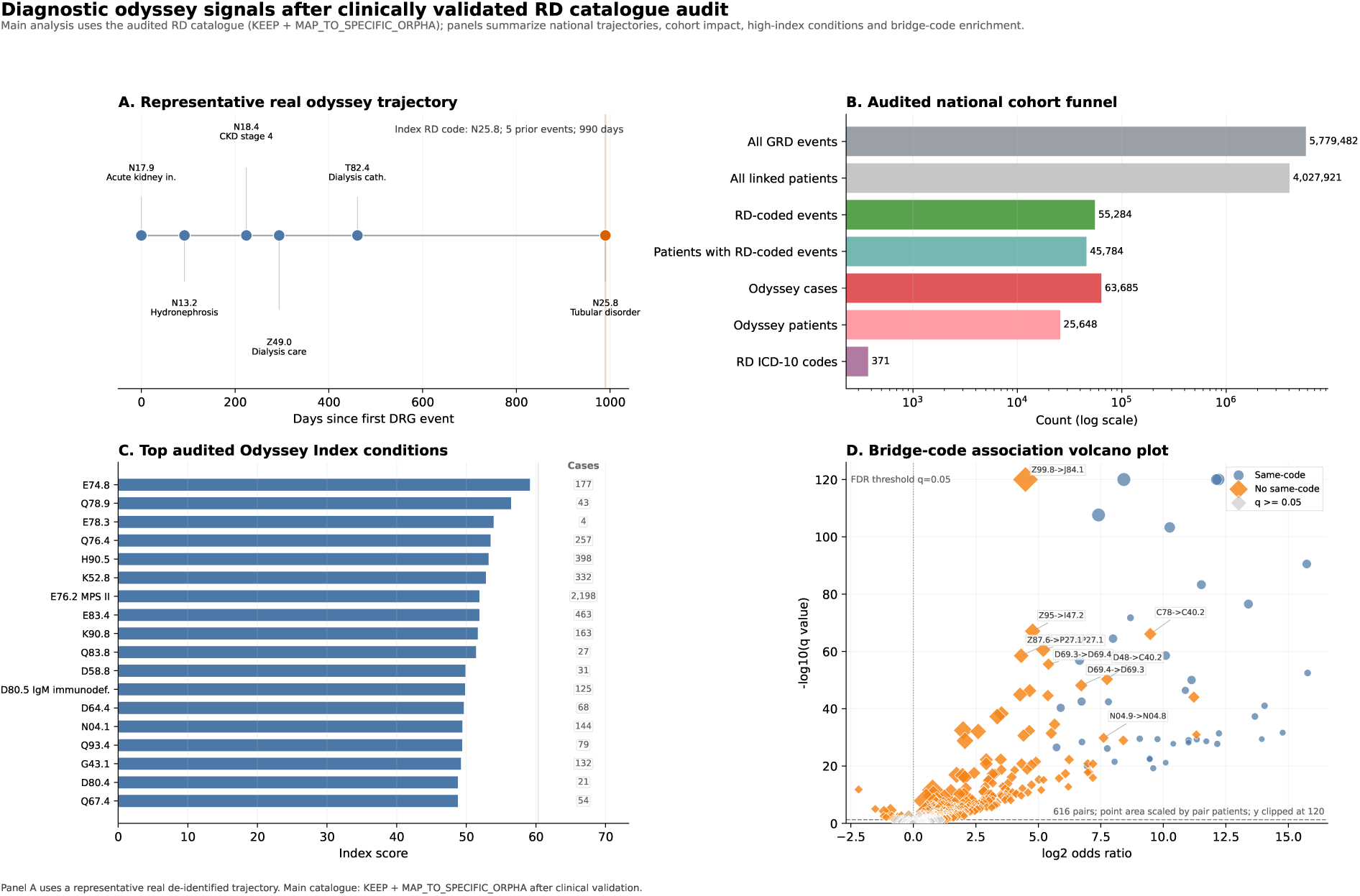
The anatomy of an inpatient diagnostic odyssey in Chilean administrative data (audited DRG/GRD-IR 2019–2024). (A) Method made visible – one real, de-identified patient. Each marker is a DRG event on the days-since-first-DRG-event axis; ICD-10 codes annotate each marker. The trajectory begins with renal/urologic and dialysis-related codes and reaches the audited rare-disease code N25.8 at the index event. Patient: 5 prior DRG events, 990 DRG-observed days to the audited N25.8 coding. **(B) Cohort funnel.** 5,779,482 GRD-IR analyzed events in 4,027,921 linked patients → 55,284 primary-diagnosis RD-coded events in 45,784 patients → 63,685 observed inpatient odyssey cases → 25,648 unique odyssey patients → 371 audited RD ICD-10 codes (catalogue: `KEEP + MAP_TO_SPECIFIC_ORPHA`, 434 codes / 701 rows). **(C) Top audited codes by Odyssey Index.** Blue bars show OI(*r*) (Eq. 1); the right-side “Cases” column reports raw odyssey-case counts for the same rows, making the separation between high-volume codes and stronger trajectory signatures explicit. **(D) Bridge-code volcano plot.** Each point is one bridge→RD-code pair with support ≥ 10 patients in the patient-unit primary analysis. The horizontal axis shows log_2_(OR), the vertical axis shows − log_10_(Benjamini–Hochberg q-value), and point area is scaled by the number of patients supporting the pair. Blue circles are same-code recurrence patterns; orange diamonds are no-same-code administrative trajectory signals. Position in the upper-right region indicates statistical enrichment and reliability under the specified model; it does not by itself establish clinical causality, diagnostic utility, or actionability. Orange no-same-code pairs were interpreted as candidate administrative trajectory signals requiring clinical review. Full estimates are reported in Table 2.

#### Output stratification

(i) Same-code pairs (bridge = RD code, recurrence). (ii) No same-code pairs (administratively distinct trajectory signals requiring clinical review).

#### Interpretation of bridge-code associations

Bridge codes are defined as the last observed primary diagnosis immediately preceding an RD-coded DRG event. They should not be interpreted as the first symptom, the first clinical manifestation, or a causal antecedent of the rare disease. Because administrative diagnosis codes combine clinical information, coding conventions, and care-process events, bridge-code associations were interpreted as statistically enriched administrative trajectory signals. We therefore considered three non-mutually exclusive interpretive classes: (i) clinically plausible precursor or manifestation, where the bridge may reflect an earlier presentation of the underlying condition; (ii) diagnostic refinement, where a broad or unspecified code precedes a more specific RD code; and (iii) care-process bridge, where the preceding code reflects treatment, follow-up, or hospitalization workflow rather than disease biology. This classification is intended to guide clinical review and prospective validation.

#### Statistical note

For each bridge-code pair, we constructed patient-level 2 × 2 tables and estimated enrichment using odds ratios, Fisher exact tests, and Benjamini–Hochberg false-discovery correction. Fisher exact testing was chosen because rare-disease data are sparse and unbalanced. The Benjamini–Hochberg procedure was used to reduce false discoveries across multiple tested bridge–RD pairs. We required support from at least 10 unique patients to reduce instability from very small counts, and applied Haldane–Anscombe correction when zero cells were present. Odds ratios are reported as measures of statistical enrichment, not as estimates of clinical effect size.

### 3.8 Odyssey Index

We propose a composite, volume-robust index that aggregates per-entity odyssey signals into a single score in [0, 100]. The index is defined per RD entity *r* for which at least three audited odyssey cases exist (*n_r_*≥ 3).

Let 𝒪*_r_*= {*i* : *c_i_* = *r, i* is an odyssey case} denote the set of audited odyssey cases coded as entity *r*, and define four per-entity summary statistics:

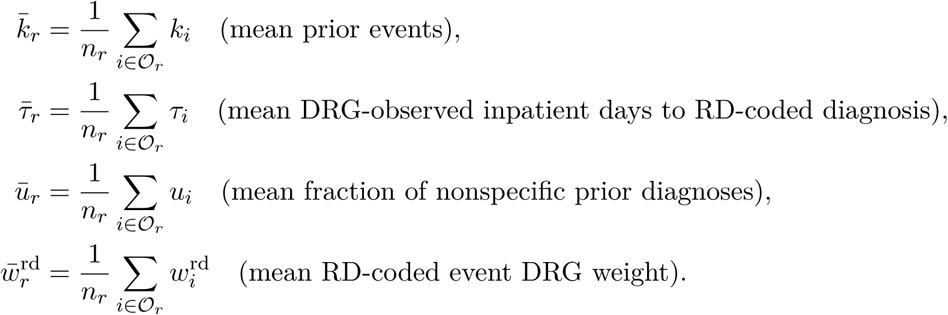

Each statistic is normalized to [0, 1] via percentile rank across the set of audited entities R:

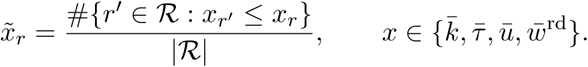

Percentile-rank normalization is preferred over min–max because it is robust to extreme-volume outliers (e.g., MPS Type II / E76.2) that would otherwise compress the rest of the distribution.

The Odyssey Index is then the weighted aggregation:

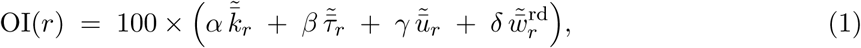

with weights *α* + *β* + *γ* + *δ* = 1. In the primary specification we use *equal weights α* = *β* = *γ* = *δ* = 0.25; sensitivity analyses re-compute OI with (i) clinically-weighted scheme (*α, β, γ, δ*) = (0.30, 0.30, 0.25, 0.15) that emphasises prior events and time, and (ii) resource-use-weighted scheme (0.20, 0.20, 0.20, 0.40). The ranking of the top-20 entities is robust to weight choice (Spearman *ρ >* 0.9 across schemes; supplement).

#### Properties

(1) OI(*r*) ∈ [0, 100] by construction. (2) The percentile-rank step makes the index *invariant to monotonic transformations* of any single component, which is desirable because DRG weight and counts are on different scales. (3) High volume alone does not push an entity to the top: an entity that has many cases but each case has few prior events will have low 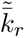. In the updated cohort, E76.2 is high volume (*n* = 2,198, 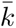 = 50.9), whereas lower-volume codes can surface when the time and nonspecificity profile is proportionally stronger.

#### Robustness

For each entity we report a 95% bootstrap CI for OI(*r*) over 1,000 case-level resamples. Entities are also re-ranked excluding the highest-volume condition to confirm the top-20 ordering does not depend on a single outlier.

### 3.9 Resource-use complexity estimation

Pre-RD cumulative DRG weight per patient and per condition is reported as a proxy for resource-use complexity; monetary conversion is avoided given the proxy nature of IR_29301_PESO.

### 3.10 Sensitivity analyses

(i) Audited conservative vs. sensitivity_no_exclude. (ii) Inclusion of secondary diagnoses. (iii) Pediatric subset. (iv) Anonymized facility-level analysis of a tertiary pediatric referral hospital in the Santiago Metropolitan Region.

### 3.11 Reproducibility

All scripts, catalogues, and templates are available in `auditoria_catalogo_epof/`. Main figures are regenerated with `regenerar_figura_principal_auditada.py`. The public GitHub repository for this paper contains the manuscript source, scripts, audited catalogues, aggregate association tables, and figure assets [19].

### 3.12 Ethics

This study used anonymized, publicly available data from the FONASA–DRG database, which contains no identifiable or individual-level personal information. All analyses were conducted using aggregated, non-identifiable records. The study protocol was reviewed by the *Scientific Ethics Committee of the Institute of Science and Innovation in Medicine (ICIM), Faculty of Medicine, Universidad del Desarrollo, Clínica Alemana*, which determined that the use of these de-identified public data did not require formal ethics approval and therefore granted an exemption.

## 4 Results

### 4.1 Cohort and final catalogue

The canonical GRD-IR source contained 5,792,715 records. After restricting by parseable admission year to 2019–2024, the analytical cohort included **5,779,482 DRG events in 4,027,921 linked patients** from 72 reporting public establishments, corresponding to FONASA-covered persons and episodes. A facility-code audit found no private clinics with observed events in this analytical cohort, despite their presence in the establishment dictionary. The Orphanet–ICD-10 homologation contained 2,247 diseases and 642 unique ICD-10 codes; the audited conservative catalogue retained 434 codes. Pre/post audit impact is shown in Table 1.

**Table 1.** National impact of audit (audited_conservative scenario vs. original).

| Metric | Original | Audited (conservative) | Reduction |
| --- | --- | --- | --- |
| Total DRG events analyzed | 5,779,482 | 5,779,482 | – |
| Total linked patients | 4,027,921 | 4,027,921 | – |
| Primary-diagnosis RD-coded events | 101,560 | 55,284 | 45.6% |
| Patients with primary-diagnosis RD-coded events | 86,404 | 45,784 | 47.0% |
| RD-coded events in any diagnostic field | 632,417 | 374,866 | 40.7% |
| Odyssey cases | 102,790 | 63,685 | 38.0% |
| Odyssey patients | 36,720 | 25,648 | 30.2% |
| Unique RD ICD-10 codes with odyssey cases | 550 | 371 | – |
| Catalogue codes / rows | 642 / 2,247 | 434 / 701 | – |

### 4.2 National magnitude of the diagnostic odyssey

Audited conservative: 55,284 primary-diagnosis RD-coded events in 45,784 patients, plus 374,866 RD-coded events in any diagnostic field. The odyssey definition yielded **63,685 observed inpatient odyssey cases in 25,648 unique patients** spanning 371 audited RD ICD-10 codes. Median DRG-observed inpatient trajectory time to RD-coded diagnosis was 241 days; mean prior events per odyssey was 8.1. This estimate is a conservative lower bound on the full diagnostic odyssey because outpatient, primary-care, emergency-without-admission, private outpatient trajectories, and private-sector FONASA hospital activity are not captured.

### 4.3 Top-20 by Odyssey Index

Figure 2, Panel C, ranks the top audited ICD-10 codes by OI(*r*) (Eq. 1). Blue bars show OI on the lower axis, while the right-side “Cases” column reports the raw number of odyssey cases for each code. This separates the trajectory-strength score from sheer volume. In the updated 2019–2024 cohort, high-index codes include E74.8, Q78.9, E78.3, Q76.4 and H90.5. E76.2 remains high volume (*n* = 2,198 odyssey cases; 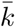 = 50.9 prior events), but its index is moderated by the combined time and nonspecificity profile; conversely, lower-volume codes such as E78.3 (*n* = 4) can surface when their trajectory profile is proportionally stronger.

### 4.4 Audited bridge codes

Of 63,685 audited odyssey cases (25,648 unique patients), 58,338 cases had a non-empty bridge code for association testing. Patient-level analysis identified **616 associations with support** ≥ 10 **patients**, **390 with** *q <* 0.05 (Fisher exact + Benjamini–Hochberg); of these, **350 were no-same-code** administrative trajectory signals. Top no-same-code associations are summarized in Table 2 and visualized in Figure 2, Panel D.

**Table 2.** Top-20 no-same-code bridge-code signals ranked by patient-level odds ratio. Audited cohort (63,685 cases, 25,648 unique patients). All listed pairs satisfy *q <* 0.05 with support ≥ 10 unique patients. CI95 are Haldane–Anscombe-corrected. Odds ratios are enrichment indicators rather than proportional estimates of clinical risk.

| RD code | Bridge | $n$ | OR | CI95 | $q$ | Representative mapping label |
| --- | --- | --- | --- | --- | --- | --- |
| E76.2 | E76.3 | 13 | 2,556 | 645–10,124 | 1.1e-31 | Mucopolysaccharidosis family |
| P27.1 | P07.2 | 27 | 2,383 | 145–39,077 | 9.1e-45 | Bronchopulmonary dysplasia |
| C40.2 | C78 | 36 | 713 | 289–1,762 | 8.4e-67 | C40.2 bone-tumour mapping |
| P27.1 | H35.1 | 19 | 338 | 91–1,260 | 1.2e-29 | Bronchopulmonary dysplasia |
| C40.2 | D48 | 32 | 215 | 116–398 | 5.1e-51 | C40.2 bone-tumour mapping |
| N04.8 | N04.9 | 20 | 195 | 71–539 | 1.5e-30 | Pierson / nephrotic-syndrome mapping |
| C40.2 | Y43.3 | 14 | 145 | 64–333 | 1.6e-21 | C40.2 bone-tumour mapping |
| P27.1 | P61.2 | 12 | 145 | 44–477 | 1.2e-16 | Bronchopulmonary dysplasia |
| Q17.2 | Q87 | 11 | 131 | 58–296 | 1.3e-18 | Microtia |
| Q28.2 | I67.1 | 14 | 126 | 48–332 | 1.5e-21 | Cerebral arteriovenous malformation |
| I45.6 | I47.1 | 11 | 126 | 57–279 | 1.6e-18 | Pre-excitation / arrhythmia mapping |
| D69.3 | D69.4 | 36 | 105 | 52–213 | 7.4e-49 | Immune thrombocytopenia / macrothrombocytopenia |
| N13.5 | Z96 | 17 | 75 | 38–150 | 5.5e-23 | IgG4-related retroperitoneal fibrosis |
| Q25.1 | P29.3 | 10 | 73 | 32–171 | 1.8e-13 | Aortic coarctation mapping |
| I73.8 | I77.1 | 16 | 68 | 27–168 | 4.4e-18 | Primary erythromelalgia / vascular mapping |
| C40.2 | Z89.6 | 13 | 57 | 29–113 | 2.0e-16 | C40.2 bone-tumour mapping |
| C40.2 | R11 | 30 | 50 | 32–79 | 2.7e-35 | C40.2 bone-tumour mapping |
| C40.2 | F43.2 | 28 | 46 | 29–73 | 3.7e-32 | C40.2 bone-tumour mapping |
| D69.4 | D69.3 | 29 | 42 | 30–60 | 2.8e-56 | Macrothrombocytopenia / thrombocytopenia |
| G80.8 | Z93.1 | 29 | 42 | 29–61 | 2.6e-45 | Worster–Drought syndrome mapping |

**Table 3.** Interpretive classes of selected bridge-code signals. Bridge-code associations may represent clinically plausible precursors, diagnostic refinement, or care-process sequencing. This classification is descriptive and intended to guide prospective clinical validation.

| Bridge pattern | Example | Likely interpretation | Strength of clinical claim | Suggested next validation step |
| --- | --- | --- | --- | --- |
| Acute organ event → chronic/inherited condition | K85.x → K86.1 | Clinically plausible precursor or manifestation | Moderate; candidate for validation | Chart review / recurrent pancreatitis pathway |
| Unspecified syndrome → specific RD code | N04.9 → N04.8 | Diagnostic refinement / coding resolution | Low to moderate | Nephrology review / biopsy-genetic-test pathway |
| Treatment encounter → RD-coded malignancy | Z51.1 → C40.2 | Care-process bridge / sequencing artifact | Low unless clinically reviewed | Coding-sequence audit |
| Related disease-family code → broader/specific RD code | E84.0/E84.8 → E84.9 | Coding-family transition | Low to moderate | Coding-family audit |
| Neurodevelopmental / seizure coding → specific RD mapping | F82/F84.0/G40.9<br>R62.x → F84.2 | Plausible diagnostic refinement, case-based | Moderate; requires validation | Neurology/genetics chart review |

#### Targeted C40.2 audit

Because C40.2 contributed multiple high-enrichment bridge-code signals, we performed a targeted aggregate audit to distinguish a possible rare bone-tumour trajectory signal from repeated oncology-care coding, referral concentration or over-specific interpretation of an ICD-10 proxy. Across the 2019–2024 DRG/GRD-IR source, C40.2 appeared in 2,563 events among 581 patients; 712 events in 377 patients used C40.2 as the primary diagnosis. Repetition and institutional concentration were substantial: 246 patients had more than one C40.2-coded event, the canonical odyssey output contained 2,025 C40.2 odyssey cases in 272 patients (7.4 cases per odyssey patient), and the leading facility accounted for 22.2% of first observed C40.2 patients but 62.5% of C40.2-coded events. Among patients with a prior DRG event before first observed C40.2, the most common immediately preceding primary diagnoses were D48 (neoplasm of uncertain or unknown behaviour; 47 patients), Z51.1 (chemotherapy session; 18 patients), Z51.8 (other specified medical care; 12 patients), and C49.2 (malignant neoplasm of connective and soft tissue of lower limb; 11 patients). We therefore retain C40.2 as an enriched administrative signal, but interpret C40.2 examples as bone-tumour coding/care-process candidates requiring chart-level validation, not as evidence of a Chilean geographic cluster or as validated adamantinoma-specific diagnostic pathways.

#### Neonatal caution for P27.1

Some catalogue-defined RD signals require special clinical caution. For example, bronchopulmonary dysplasia is represented in rare-disease terminologies and appears in the audited catalogue, but in clinical practice it is often understood as a complication of prematurity, mechanical ventilation and oxygen exposure rather than as a prototypical monogenic rare disease. We therefore interpret P27.1 bridge-code patterns as catalogue-defined administrative RD signals requiring neonatal clinical review, not as individually validated rare-disease diagnoses.

Because rare-disease bridge-code pairs are sparse, very large odds ratios should be read as enrichment indicators rather than as proportional estimates of clinical risk. We therefore interpret the table jointly using effect size, q-value, support, and clinical-administrative plausibility.

### 4.5 Pre-diagnostic resource-use complexity

Cumulative pre-RD DRG weight per condition is reported in the supplement (per ICD-10 chapter and per top entity). Mean DRG weight of the RD-coded event ranges from ≈ 0.5 to ≈ 2.1 across the leading entities.

### 4.6 Illustrative case: the anatomy of a renal/tubular odyssey

#### Pre-index renal trajectory (days 0–461)

Figure 2, Panel A, displays the inpatient trajectory of a single, real, de-identified patient whose first observed audited RD endpoint is **N25.8**. In the audited catalogue, N25.8 is retained as a clinically reviewed renal tubular-disorder code, mapped to rare tubulopathies such as Dent disease and pseudohypoaldosteronism type 1. Each marker on the time-line is one coded DRG event; the *x*-axis is days since the first DRG event of this patient, and the orange marker around day 990 is the index event carrying the audited RD code after five prior observed events.

The preceding DRG events move from acute kidney injury (N17.9) and hydronephrosis/urolithiasis coding (N13.2) to chronic kidney disease stage 4 (N18.4), dialysis-related care (Z49.0) and a dialysis-catheter complication (T82.4). The sequence is clinically coherent as an administrative renal trajectory, but it does not by itself establish causality or a missed diagnosis.

#### Index event (day 990)

The audited RD code N25.8 appears at the index event after five prior DRG records and 990 DRG-observed days. This is the point at which the pipeline closes the observed inpatient odyssey for this patient-level trajectory.

#### Why this case is informative

The case is deliberately used as a transparent, non-synthetic example of the pipeline rather than as clinical proof. It shows how repeated organ-system-specific hospital coding can precede a later audited RD code by months to years. As with all DRG-based odyssey measures, the outpatient portion of the diagnostic history is invisible; the reported 990 days are therefore a lower bound on the real diagnostic delay (cf. §3.2).

### 4.7 Facility-level feasibility analysis: anonymized pediatric use case

To illustrate the operational translation of the national pipeline, we applied the audited RD catalogue and odyssey definitions to an anonymized tertiary pediatric referral hospital in the Santiago Metropolitan Region. This analysis is intended as a facility-level feasibility use case and should not be interpreted as a formal institutional performance assessment. Under the updated canonical DRG/GRD-IR 2019–2024 source, the facility contributed **524 primary-diagnosis RD-coded events in 395 patients**, equivalent to **2.8%** of its DRG events. The broader facility-associated odyssey definition identified **231 cases in 154 patients** (**12.3 cases per 1,000 DRG events**), while the stricter first-late-RD subset identified **101 episodes in 97 patients**, with a median observed inpatient time to first late RD coding of **216 days**. Supplementary Figure S3 illustrates how the audited catalogue, odyssey rate, leading RD diagnoses, and strict first-late-RD subset can be operationalized at facility level.

## 5 Discussion

### Central finding

The RD diagnostic odyssey in Chile is detectable even through the restricted DRG-observed window. Using an audited Orphanet–ICD-10 catalogue, we identified a substantial cohort of patients with prior DRG events before an RD-coded event, quantified DRG-observed inpatient trajectory time, and detected statistically enriched bridge-code patterns. Because DRG/GRD-IR data exclude primary care, outpatient specialty care, emergency encounters without admission, private outpatient trajectories, private-sector FONASA hospital activity, laboratory results, imaging, and genetic testing, these estimates represent a conservative lower bound on the full diagnostic odyssey. The main contribution of this work is therefore not the clinical validation of individual bridge-code pairs, but the development of a reproducible national infrastructure for RD trajectory surveillance and prioritization.

### Comparison with Avila 2022

Explicit complementarity: they quantified *mortality* (final outcome, DEIS 2002–2017); we describe *morbidity and inpatient trajectory* (process, DRG/GRD-IR 2019–2024).

### Methodological validity

Auditing reduces primary-diagnosis RD-coded events by 45.6% relative to the raw catalogue, evidencing that prior analyses based on un-audited homologations may inflate rates. Comparison with the sensitivity_no_exclude scenario provides a plausible range around the conservative primary estimates.

### Positioning vs. agentic diagnostic systems

ThinkRare [6] and DeepRare [7] operate at the *individual* level (find the patient, suggest the diagnosis). Our pipeline operates at the *population/regulatory* level: it defines and audits the entities prior to any analysis and characterizes the trajectory at the system scale. The two layers are complementary.

### Clinical and regulatory implications

Clinically plausible bridge-code signals, such as acute pancreatitis preceding hereditary chronic pancreatitis or unspecified nephrotic syndrome preceding specific nephrotic-syndrome mappings, may be prioritized for prospective review in sentinel hospitals. These signals are not ready-to-use alert rules; rather, they provide a reproducible evidence base for designing and evaluating future referral, coding-quality, or clinical-decision-support workflows. In regulatory terms, these findings should be interpreted as policy-relevant evidence rather than as a formal implementation instrument. They can inform the construction and revision of the national rare-disease list under Law 21,743 (Art. 5), support the analytic and quality-control layer of the National Registry (Art. 6, with the technical norm already in force), and provide a reproducible evidence-generation framework for the Technical Advisory Commission (Art. 4). They may also complement Law 20,850 (Ricarte Soto) coverage monitoring and Line 1 of the proposed National Plan.

### Latin American generalizability

The pipeline is applicable to any system with administrative ICD-10 (Colombia, Argentina, Mexico, Peru); the agentic skill is reusable. Regional adoption would contribute to the *Rare 2030* foresight study and to the International Rare Diseases Research Consortium (IRDiRC) goal of diagnosis ≤ 1 year.

### Future directions

Future work should integrate pharmacovigilance and genetic testing data; prospectively validate candidate bridge-code signals in sentinel hospitals; integrate DEIS-MINSAL mortality to connect trajectory and outcome; link DRG/GRD-IR with primary care, ambulatory emergency, FONASA claims, and private-sector records to overcome the current DRG-observed window and reconstruct the full odyssey; and adapt the audited catalogue to future ICD-11 implementation. The transition to ICD-11 is particularly relevant for rare diseases because its improved semantic structure and close relationship with rare-disease terminologies such as Orphanet may reduce ambiguity inherent to ICD-10-based ascertainment [17, 18]. Agentic diagnostic copilots such as DeepRare and ThinkRare could then be anchored on better-governed, interoperable RD catalogues.

## 6 Limitations

### 6.1 Structural limitations of DRG data (most important)

- **DRG events only.** The GRD-IR source captures hospital discharges and major ambulatory surgery from reporting public establishments for FONASA-covered persons, but does not capture outpatient specialty care, primary care, emergency without admission, private outpatient care, or private-sector hospital activity absent from the observed files. The diagnostic odyssey occurring before the first DRG-observed event is invisible. By construction, all reported times and counts are a *conservative lower bound on the total diagnostic odyssey*.
- **No observed private-sector FONASA activity.** Although the establishment dictionary includes private clinics, the 2019–2024 analytical cohort contained no observed events from private clinics such as Clínica Alemana, Clínica Las Condes, or Clínica Santa María. This implies that FONASA-covered activity occurring through private-sector mechanisms, including *Modalidad Libre Elección* or *Ley de Urgencia*, is not represented in the present analytical cohort. Patients treated through ISAPRE/private-insurance circuits are also absent unless they later appear in the public FONASA DRG/GRD-IR data.
- **No auxiliary clinical information.** DRG data contain no laboratory results, imaging, genetic panels, sequencing, narrative epicrisis, chief complaint, or evolution.
- **No outpatient treatment.** Chronic pharmacotherapy, gene therapy, and biologics covered by Law 20,850 not leading to hospitalization are not captured; reported pre-diagnostic resource-use complexity is strictly inpatient.

### 6.2 RD operational catalogue

- ICD-10-based operational definitions underestimate very rare entities coded with broad codes; 118 `NARROW_DEFINITION` codes still lack implemented narrow rules and remain outside the primary analysis.
- 44 REVIEW_CLINICAL codes also remain outside the primary analysis until further human review.
- Primary analysis uses `DIAGNOSTICO1`; secondary diagnoses change counts and trajectories (analyzed partially as sensitivity).
- Auditing improves specificity but may reduce sensitivity for rare phenotypes coded with broad ICD-10 codes.

### 6.3 Temporal and resource-use complexity

- The 2019–2024 window includes the COVID-19 period (2020–2022) with displacement of elective hospitalizations and plausible underreporting.
- `IR_29301_PESO` is a complexity / resource-use proxy, not a monetary amount.

### 6.4 Bridge-code interpretation

- Bridge-code associations are based on the immediately preceding primary diagnosis and therefore capture the last observed administrative step before an RD-coded DRG event, not the full pre-diagnostic clinical history. Some associations may reflect clinically plausible precursor manifestations, whereas others may reflect diagnostic refinement, same-family ICD-10 transitions, treatment encounters, or hospital coding practices. The bridge-code analysis should therefore be interpreted as a hypothesis-generating prioritization layer requiring clinical review and prospective validation before use in alert systems, referral rules, or policy instruments.

### 6.5 Statistical interpretation

- Rare-disease administrative data produce sparse contingency tables, and odds ratios can become very large when bridge–RD pairs are uncommon outside a narrow diagnostic family. We therefore interpret odds ratios as enrichment measures and report them together with q-values and patient support, rather than as direct estimates of clinical risk or causal magnitude.

### 6.6 Linkage

- The anonymized patient ID supports longitudinal linkage within the FONASA DRG/GRD-IR files, but not across GRD-IR and other registries, including DEIS-MINSAL mortality records, monthly statistical summaries (*Resúmenes Estadísticos Mensuales*, REM), broader FONASA insurance and claims datasets, or private-insurance records from ISAPREs. Such integration requires agreements not feasible for this version.

## 7 Conclusion

To our knowledge, we report the first nationwide audited and reproducible characterization of inpatient RD diagnostic odysseys in Latin America, based on a clinically reviewed Orphanet–ICD-10–DRG/GRD-IR operational catalogue. The Odyssey Index and bridge-code analysis provide a quantitative substrate for trajectory surveillance, hypothesis generation, and facility-level prioritization under Chile’s Law 21,743. Bridge-code associations should be interpreted as statistically enriched administrative signals requiring prospective clinical validation before use in alert systems, referral rules, or formal policy instruments. The same framework may support national registry design, quality-control analyses, Ricarte Soto coverage monitoring, and sentinel-hospital validation studies.

## Data Availability

The data analyzed in this study are publicly available through FONASA Datos Abiertos. The public repository for this paper contains the manuscript source, analysis scripts, audited catalogues, aggregate association tables, and figure assets: https://github.com/gear-go/epof-odiseas-diagnosticas-paper. Repository folder names retain the original Spanish EPoF acronym used during pipeline development.

## Data and code availability

- Public repository: https://github.com/gear-go/epof-odiseas-diagnosticas-paper.
- Audited catalogue and per-code decisions: `auditoria_catalogo_epof/outputs/`.
- Clinical-validation template: `auditoria_catalogo_epof/evidencia/revision_clinica/`.
- Agentic skill: `auditoria_catalogo_epof/skills/validacion-clinica-epof/`.
- Audit plan and reproduction scripts: auditoria_catalogo_epof/.
- Main figure, volcano panel and companion CSVs: repository root figure outputs.
- Bridge-code association tables: output folder `bridge_code_associations/`.
- DRG/GRD-IR source files: FONASA Datos Abiertos public portal.

Repository folder names retain the original Spanish EPoF acronym used during pipeline development.

## Author contributions (CRediT, initial)

**GGV** – conceptualization, methodology, software, formal analysis, data curation, original draft, visualization. **GR** – conceptualization, clinical supervision, clinical validation of audited catalogue, critical review. **LB** – methodology (data science, trajectory mining), methodological supervision, critical review. **CC-L** – epidemiology and health policy, Chilean-system contextualization (continuity with Encina et al. 2019), critical review. **ID** – DRG/GRD-IR data management, extraction of relevant analytical information, data validation, critical review. **MIMW** – clinical/epidemiological validation, RD/EPoF policy contextualization, critical review.

## Ethics statement

This study used anonymized, publicly available data from the FONASA–DRG database, which contains no identifiable or individual-level personal information. All analyses were conducted using aggregated, non-identifiable records. The study protocol was reviewed by the Scientific Ethics Committee of the Institute of Science and Innovation in Medicine (ICIM), Faculty of Medicine, Universidad del Desarrollo, Clínica Alemana, which determined that the use of these de-identified public data did not require formal ethics approval and therefore granted an exemption. The illustrative trajectory is displayed only at the ICD-10-code and relative-time level, without calendar dates, institution sequence, sex, age, municipality, or other quasi-identifiers.

## Acknowledgements

We thank FONASA for publishing open GRD/GRD-IR data.

## Supplementary Materials

**Figure S1.**
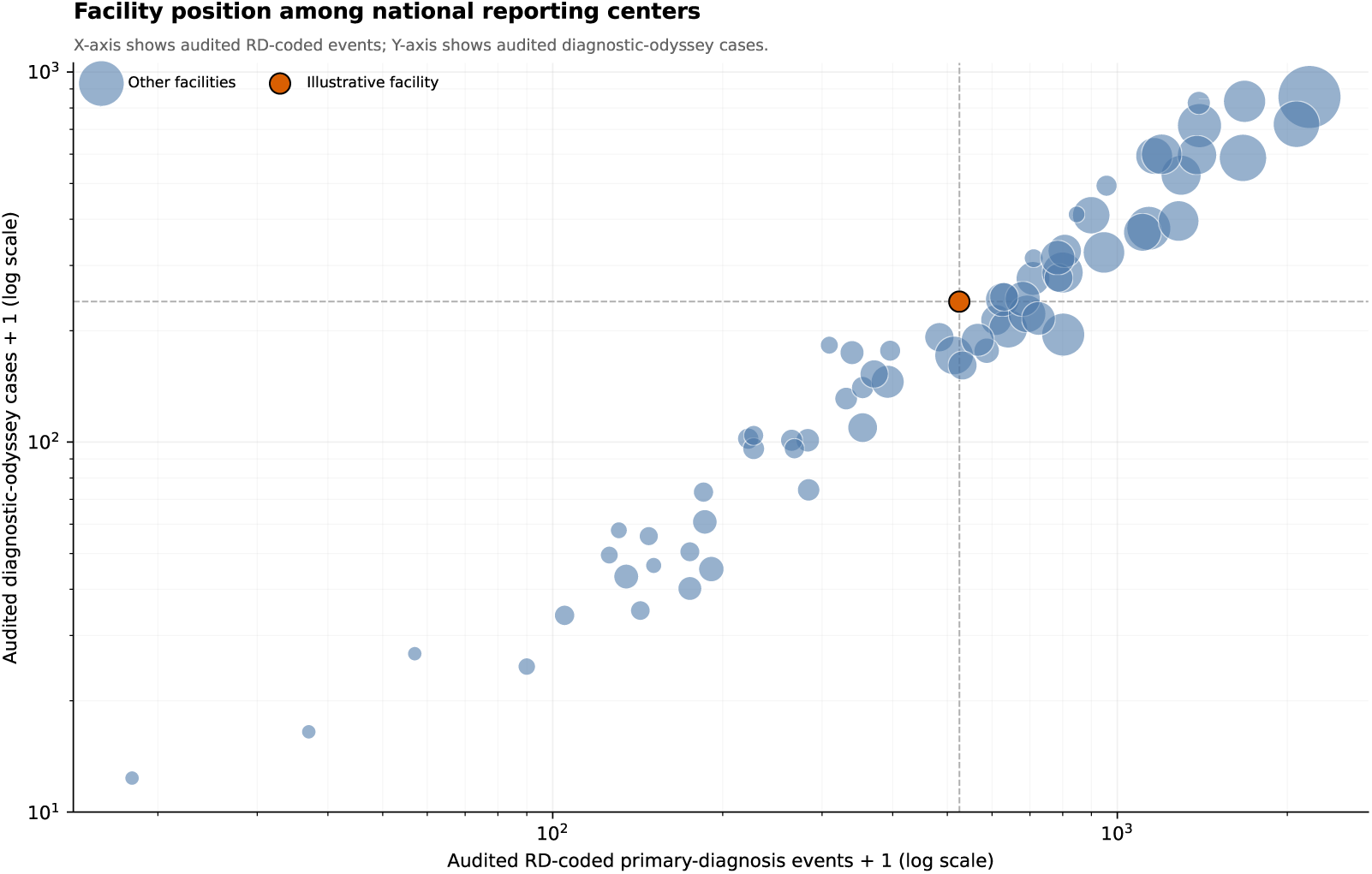
Facility-level position among national reporting centers. Each marker is a reporting facility; the horizontal axis shows audited RD-coded primary-diagnosis events, and the vertical axis shows observed diagnostic-odyssey cases. The orange marker is the anonymized illustrative pediatric facility. *Marker area* is scaled by total DRG events.

**Figure S2.**
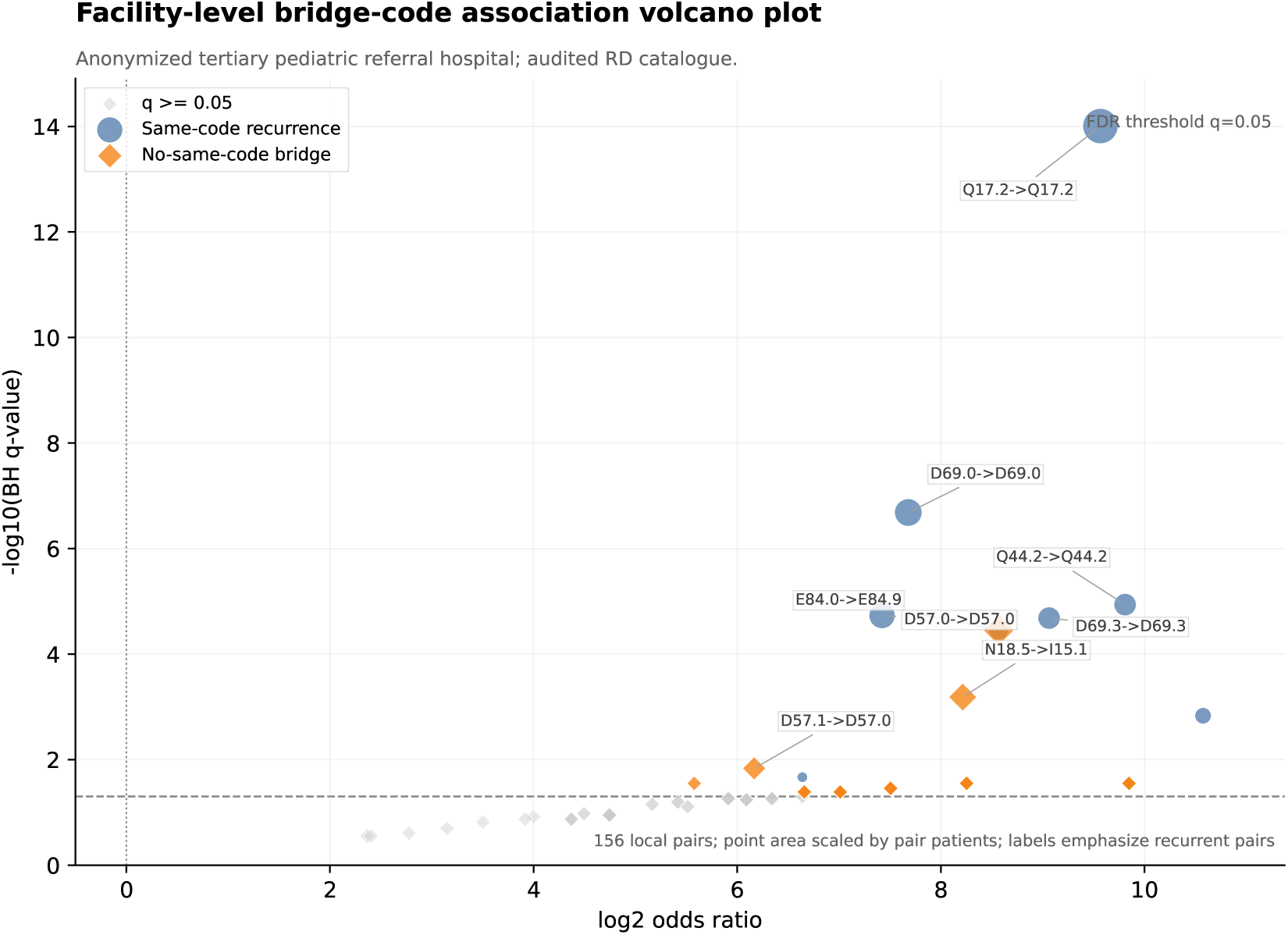
Facility-level bridge-code association volcano plot. Patient-level Fisher exact tests with Benjamini–Hochberg correction were recomputed after restricting the audited RD odyssey cohort to the anonymized tertiary pediatric referral hospital. Each point is an observed bridge→RD-code pair; the horizontal axis shows log_2_(odds ratio), the vertical axis shows − log_10_(BH q-value), and point area is proportional to the number of local patients supporting the direct pair. Blue circles are same-code recurrence patterns and orange diamonds are no-same-code bridge candidates. Singleton-supported pairs are retained as exploratory local background, while labels emphasise recurrent pairs. The local signal is dominated by expected same-code recurrence, with a small number of clinically interpretable no-same-code pairs such as E84.0→E84.9, N18.5→I15.1, and D57.1→D57.0; interpretation is exploratory because local support is necessarily lower than in the national primary analysis.

**Figure S3.**
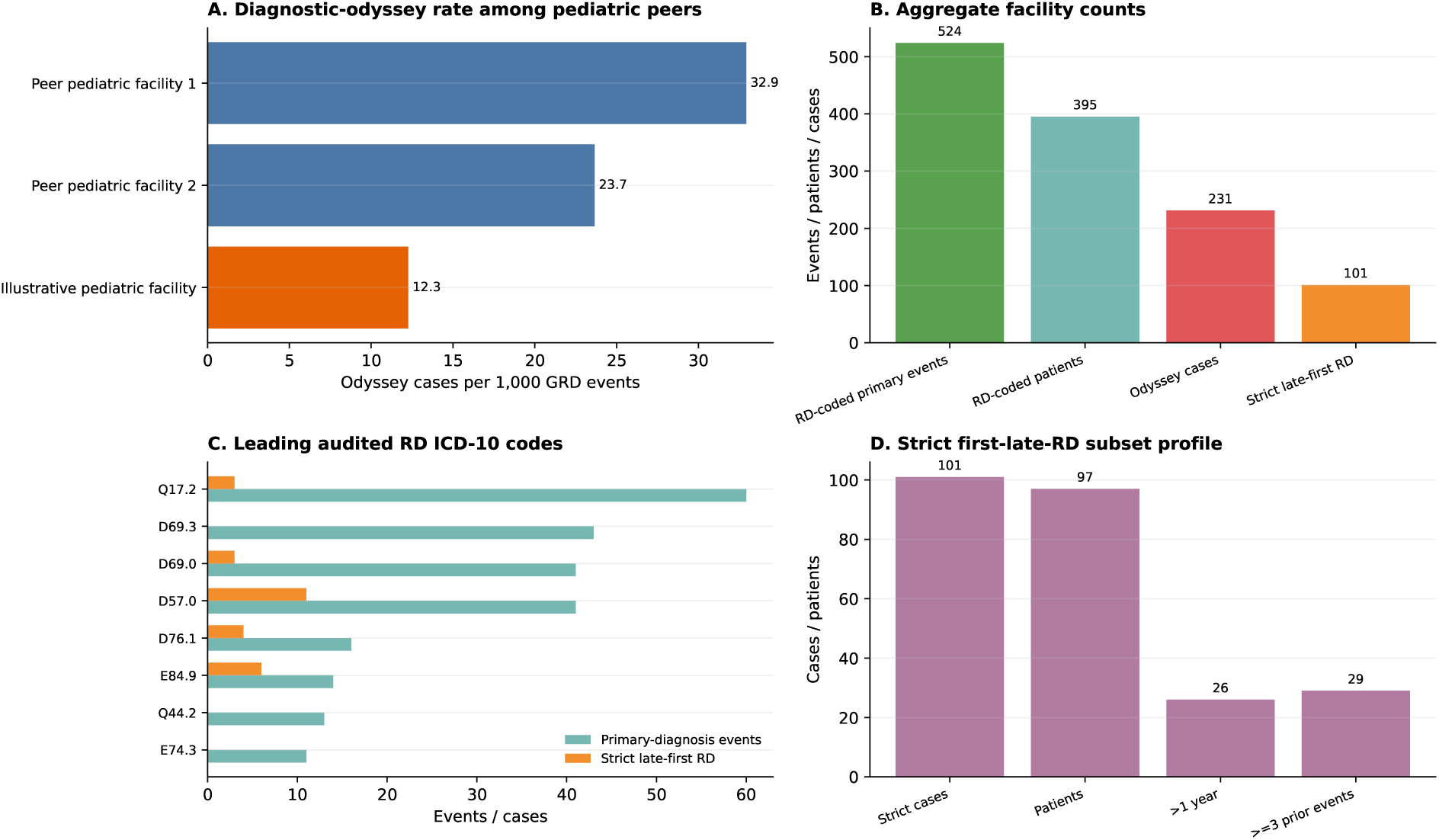
Facility-level feasibility analysis – anonymized tertiary pediatric referral hospital (audited DRG/GRD-IR 2019–2024). **(A) Diagnostic-odyssey rate among pediatric referral facilities.** The anonymized illustrative facility is shown in orange. **(B) Aggregate facility counts.** The panel summarizes audited RD-coded primary-diagnosis events, patients, observed diagnostic-odyssey cases, and the strict first-late-RD subset. **(C) Leading audited RD ICD-10 codes at the facility.** For each top code we show total audited RD-coded events (teal) and the strict first-late-RD subset (orange). **(D) Strict first-late-RD subset profile.** The panel summarizes strict cases, patients, cases with more than one year of observed inpatient time, and cases with at least three prior events.

## Abbreviations

RD: rare diseases
EPoF: *enfermedades poco frecuentes*
DRG: Diagnosis-Related Groups
GRD/GRD-IR: Chilean/Spanish DRG terminology and International Refined GRD implementation
FONASA: National Health Fund
ISAPRE: private health insurer.

